# The Proportional Treatment Effect: A Metric That Empowers and Connects

**DOI:** 10.1101/2025.02.12.25322182

**Authors:** Guoqiao Wang, Yijie Liao, Caiyan Li, Kun Jin, Yan Li, Gary Cutter

## Abstract

Clinical trials with continuous endpoints, evaluate efficacy by comparing the difference in mean changes from baseline between groups. However, clinicians often interpret results in terms of a proportional reduction rather than an absolute difference. An alternative approach is to reparametrize this difference as a proportional treatment effect, calculated by dividing the difference by the placebo mean change. We demonstrate that, in theory, the proportional treatment effect can be more powerful than the simple difference in means while still controlling the type I error rate. This is achieved using the delta method as implemented in well-established computational tools like the R package ‘msm’ and the SAS procedure ‘NLMIXED’. By analyzing data from phase III trials, we illustrate how a proportional treatment effect connects treatment outcomes across various endpoints and different presentation formats. The availability of these well-established statistical tools for estimating proportional treatment effects, combined with this theoretical demonstration, suggests an alternative test statistic for clinical trials with continuous endpoints.

## 1. Introduction

In clinical trials with continuous endpoints, efficacy inference has traditionally been based on comparing the difference (*μ*_*P*_ − *μ*_*T*_) in the mean change from baseline to the last study visit between the placebo group (*μ*_*P*_) and the treatment group (*μ*_*T*_) using a two-sample t-test. Recently, an alternative approach has been proposed, which involves reparameterizing the difference as a proportional treatment effect: 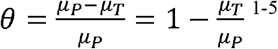. It has been demonstrated that under the same model setting, this reparameterization can increase power compared with the difference between means.^1, 2, 4^ However, this conclusion is currently based on simulations and has not yet been derived theoretically in any literature. This proportional treatment effect essentially relates to the ratio of two means, a topic that has been extensively investigated,^6-9^ including the concise confidence interval formula provided by Fieller’s theorem.^10^ Nonetheless, none of these investigations have directly explored why a proportional treatment effect can potentially have greater power than the difference between the means of the treatment and placebo groups while controlling Type I error.

In this study, we provide theoretical justification regarding the following points based on the delta method as implemented in well-established computational tools like the R package ‘msm’ and the SAS procedure ‘NLMIXED’: (i) A proportional treatment effect, under certain conditions, can lead to greater power than the difference in means (henceforth referred to as the placebo-treatment difference). (ii) A proportional treatment effect does not inflate Type I error. (iii) A proportional treatment effect connects various ways of measuring treatment efficacy across different endpoints within the same clinical trial.

## 2. Materials and Methods

A conventional clinical trial with a continuous endpoint typically features a randomized (often 1:1 ratio), placebo-controlled, double-blind design with parallel groups (or active comparator, but for simplicity we will only use placebo without loss of generality). For a given primary continuous endpoint, either higher or lower values indicate better outcomes. Let *μ*_*P*_ denote the mean change from baseline for the placebo group, and *μ*_*T*_ for the treatment group. Without loss of generality, we assume that the investigational drug leads to improvement in the endpoint. That means *μ*_*P*_ ≤ *μ*_*T*_ < 0 when higher values indicating better outcomes; and *μ*_*P*_ ≥ *μ*_*T*_ > 0 when lower values indicating better outcomes. We define the proportional treatment effect as: 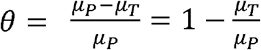. Therefore, regardless of the direction (negative or positive) of the mean change from baseline, *θ* is positive (i.e., *θ* > 0) when the treatment effectively slows down the disease progression.

The test statistic for the difference in the mean change from baseline between the placebo and treatment groups was compared with the test statistic for a proportional treatment effect under this conventional clinical trial setting. The connection between various measures of the treatment effect, and between the treatment effects across different endpoints, are demonstrated using published data from the Clarity AD^11^ and TRAILBLAZER-ALZ2^12^ trials for Alzheimer’s disease.

## 3. Results

### 3.1 A Proportional Treatment Effect Empowers and Controls Type I Error

Let us consider a two-sample situation where the random variables are assumed to be mutually independent and normally distributed with an unknown but common variance *σ*^2^. Specifically, let *X*_*jP*_ ∼ *N*(*μ*_*P*_, *σ*^2^) be the *N* observations of the change from baseline to the last visit in the placebo group and *X*_*jT*_ ∼ *N*(*μ*_*T*_, *σ*^2^) be the *N* observations in the treatment group with *j* = 1, … , *N. Without loss of generality, we assume that lower values indicating better outcomes and μ*_*P*_ ≥ *μ*_*T*_. Traditionally, the efficacy inference is based on the difference between the two group means. Let 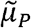 be the mean of these *X*_*jP*_ observations, then 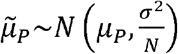. Similarly, 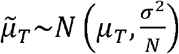. The test statistic of the difference *μ*_*P*_ − *μ*_*T*_ can be estimated as:

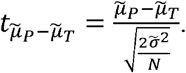

When the proportional treatment effect is defined as:

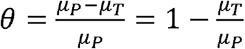 then *μ*_*T*_ = *μ*_*P*_ (1 − *θ*) Subsequently, *X*_*jP*_ ∼ *N*(*μ*_*P*_, (1 − *θ*) *σ*^2^).

By the classical delta method,^10^

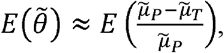

and, furthermore, *X*_*jT*_ is independent of *X*_*jP*_, thus

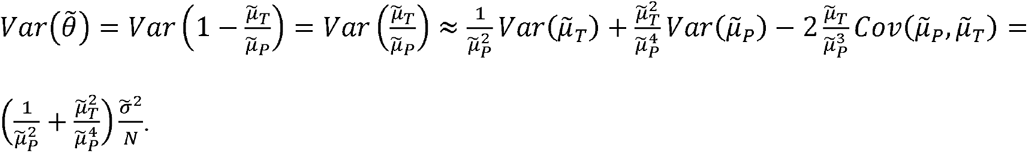

It is worth noting that the use of this formula (i.e., first-order derivative) to approximate the variance of a ratio has been widely implemented across various studies^6, 9, 13, 14^ and statistical packages, including R packages “msm”^15^ and “car”^16^, as well as SAS “proc nlmixed”.^3, 17^ Our goal is to establish a connection between the test statistic of *θ* and the test statistic of the difference by applying this well-established variance formula.

Given the variance, the test statistic of *θ* can be estimated as:

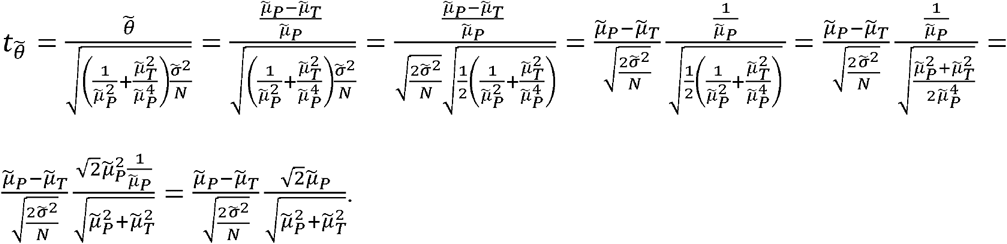

When 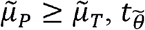 can be related to 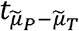 in the following way:

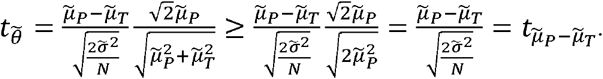

Therefore, a proportional parameterization will lead to a larger test statistic, consequently yielding a smaller p-value and greater power. Under the null hypothesis, when *μ*_*P*_ = *μ*_*T*_, we have 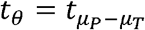. Therefore, both test statistics result in the same type I error control.

It is worth noting that the power gain might be attributed to the delta method, which is used to approximate the distribution of the test statistic. Under these circumstances, our derivation reveals a potential limitation in commonly used statistical software packages when estimating nonlinear proportional treatment effects. It is crucial for statisticians to be aware of this issue. To the best of our knowledge, this matter has not been addressed in the existing literature.

### 3.2 Reparameterization Matters

When *μ*_*P*_ ≥ *μ*_*T*_ and 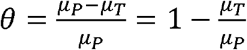, Section 3.1 demonstrates that this proportional treatment effect can have greater power than the difference. However, when the proportional treatment effect is defined differently as: 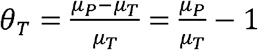. The subscript *T* in *θ*_*T*_ indicates the proportional treatment effect is relative to the mean of the treatment group rather than the mean of the placebo group as in *θ*. Following the same derivation process described in Section 3.1, it can be showed that:

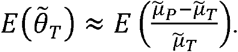

And,

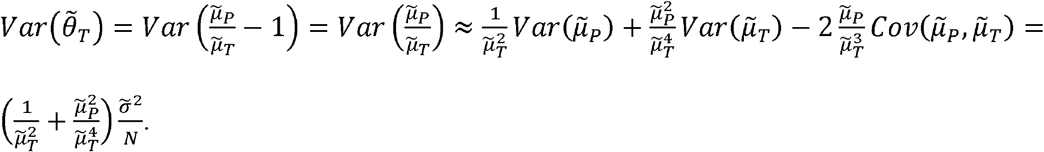

Therefore, the test statistic of *θ*_*T*_ can be estimated as:

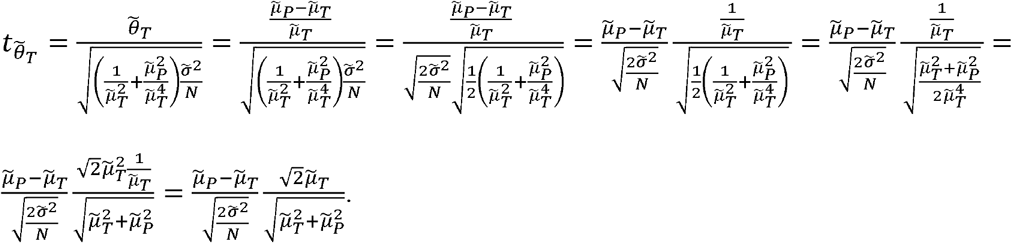

Similarly, when 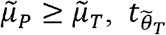 can be related to 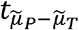 in the following way:

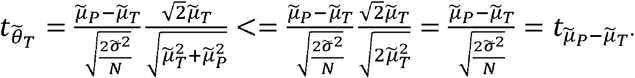

Therefore, this proportional parameterization will lead to a smaller test statistic, consequently yielding a larger p-value and less power. Under the null hypothesis, when 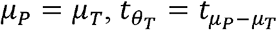. Therefore, the type I error is still controlled.

### 3.3 Simulation Results

To illustrate the theoretical conclusions presented in Sections 3.1 and 3.2, simple simulations were performed to model a two-sample scenario as described in Section 3.1. Three test statistics were evaluated: (i) the proportional effect with the larger mean as the denominator 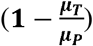; (ii) the proportional effect with the smaller mean as the denominator 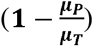; and (iii) the difference between the two means (*μ*_*P*_ – *μ*_*T*_). Table 1 presents the simulation results, which were generated using the SAS procedure *nlmixed* (supplemental material includes the corresponding SAS code).

**Table 1:** Power and Type I error for various simulated scenarios.

| Method | Power | | $\mu_T = 0.9$ | Type I Error<br>$\mu_T = 1$ |
| --- | --- | --- | --- | --- |
| | $\mu_T = 0.7$ | $\mu_T = 0.8$ | | |
| Proportional Effect ($1 - \frac{\mu_T}{\mu_P}$) | 63.7% | 38.9% | 13.9% | 3.9% |
| Proportional Effect ($1 - \frac{\mu_P}{\mu_T}$) | 33.3% | 15.7% | 10.3% | 4.5% |
| Difference ($\mu_P - \mu_T$) | 55.5% | 30.1% | 4.6% | 4.1% |
| $\mu_P = 1, SD = 1$<br>Two-sided t-test with type I error of 5% | | | | |

These results validate the theoretical conclusions, demonstrating that estimating the nonlinear model with the standard SAS procedure can reveal that the proportional treatment effect offers greater power than the traditional difference while maintaining Type I error control. Furthermore, the proportional treatment effect exhibits greater power when the larger mean is used as the denominator. In other words, reparameterization is critical.

### 3.4 A Proportional Treatment Effect Connects

For clinical trials with a continuous primary endpoint, such as the Clarity AD^11^ and TRAILBLAZER-ALZ2^12^ trials for Alzheimer’s disease, various efficacy measures have been used to present the treatment effect, including the difference in the mean change from baseline between groups, time savings in disease progression, and reduction in hazard ratio. Despite these diverse representations of the treatment effect, when converted to a proportional treatment effect, they converge to very similar values within the same trial (Table 2).

**Table 2:** Illustration of various efficacy measures and their interconnections.

| Study | Efficacy Measures | Efficacy Result | Proportional Treatment Effect* |
| --- | --- | --- | --- |
| Clarity AD | Difference in mean change<sup>□</sup> | 0.45 | 27% = 0.45/1.66 |
|  | Time savings (months)<sup>□</sup> | 5.3 | 29% = 5.3/18 |
|  | Hazard ratio<sup>□</sup> | 69% | 31% |
| TRAILBLAZER-ALZ2<br>(Low/Median Tau Population) | Difference in mean change | 0.67 | 36% = 0.67/1.88 |
|  | Time savings (months) | 7.0 | 40% = 7 × 52/12/76 |
|  | Hazard ratio | 61% | 39% |
| <sup>□</sup> These measures were estimated based on CDR-SB score<br><sup>□</sup> Hazard ratio was estimated based on progression in CDR Global score<br>*Proportional reduction relative to the placebo group. For example, when the hazard ratio of treatment to placebo is 69%, the relative reduction is 31%. |  |  |  |

Additionally, each clinical trial typically employs multiple key secondary endpoints with various scale ranges. Comparing the treatment effects (i.e., the placebo-treatment difference) across primary and secondary endpoints can be challenging due to these differing scales. Figure 1 illustrates the comparison of two different representations of treatment effects for both the Clarity AD and TRAILBLAZER-ALZ2 (low/median tau population) trials. Despite the large variation in the differences between means for the four endpoints in each trial, converting these to proportional treatment effects (*θ*) relative to the placebo mean change makes the treatment effects more comparable and easier to interpret, both within the same trial and across trials. Table 2 and Figure 1 demonstrate that using a proportional treatment effect not only unifies various efficacy measures but also aligns treatment effects across different endpoints within the same trials.

**Figure 1:**
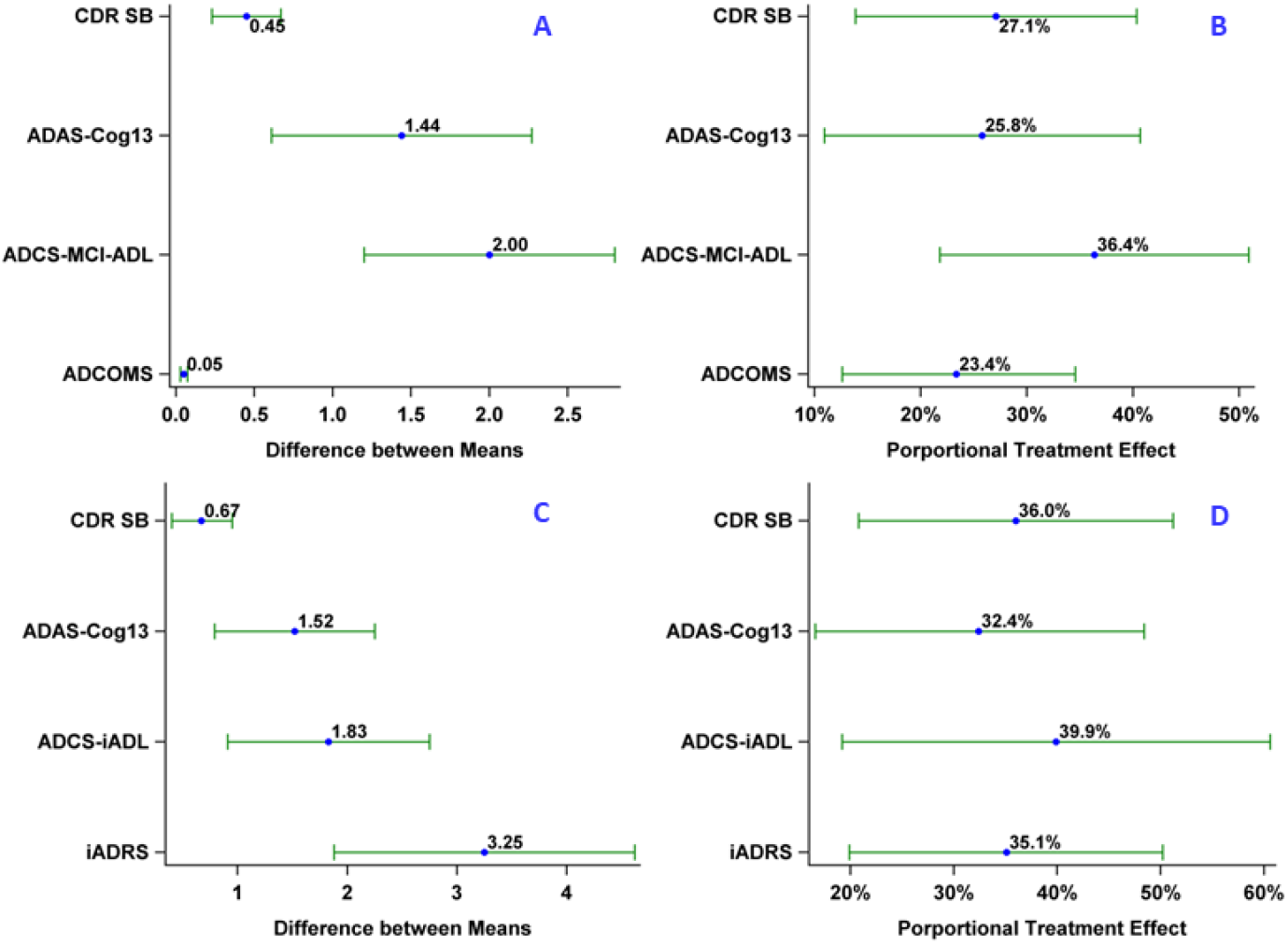
Comparison of two different types of presentation of treatment effects. Panels A and C: the difference between means; Panel B and D: the proportional treatment effect. Panels A and B were generated using data from the Clarity AD trial. Panels C and D were generated using data from the TRAILBLAZER-ALZ2 trial (low/median tau population).

### 3.5 Bias and Asymmetry

It has been shown that the distribution of 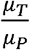 can be asymmetric, and its estimate is asymptotically unbiased with a bias given by 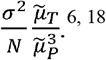. Consequently, the proportional treatment effect 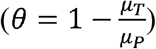 can also exhibit asymmetry, and its estimate is asymptotically unbiased. The bias is of *θ* is 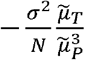. In other words, the estimate of *θ* based on the delta method provides a smaller estimate than the true *θ* when both means are either positive or negative and *θ* is positive.

## 4. Discussion

The use of a proportional treatment effect is well established in the analysis of survival time-to-event endpoints (e.g., Cox proportional model)^19^ and categorical endpoints (e.g., proportional odds ratio model^20^). A proportional effect provides a flexible way to evaluate the average treatment efficacy between groups and has become a standard tool for both survival and categorical endpoints. For continuous endpoints, efficacy inference traditionally relies on the difference in mean change from baseline between groups.^11,12^ The methodologies and computational packages needed to analyze this difference are well established. Additionally, estimating sample size when using the difference for efficacy inference is straightforward.

In contrast, although the proportional treatment effect stemming from reparameterization using the difference and the placebo mean 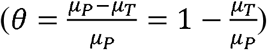 has been extensively used to communicate treatment effects, it has not been widely employed as a formal efficacy inference test statistic. For example, in the Clarity AD trial, a 27% reduction relative to the placebo decline in CDR-SB, obtained by dividing the difference between groups by the placebo mean (27% = 0.45/1.66), has been used to communicate the treatment effect to clinicians, patients, and the media. Similarly, in the TRAILBLAZER-ALZ2 trial (low/medium tau population), a 36% reduction has been widely reported. A proportional treatment effect is often easier to communicate than the difference in primary endpoints, especially for non-researchers.

In this report, we demonstrate that a proportional treatment effect can potentially have greater power under certain circumstances than the difference, even though the former is a reparameterization of the latter. Our demonstration was based on the popular nonlinear approximation method employed in various well-established computational packages. We also show how reparameterization matters. Because the variance of the proportion is a function of the ratio of the two means, to reduce the variance, the larger mean should be the denominator. Furthermore, we illustrate that regardless of how treatment effects are presented, and which endpoint is used, they are interconnected once converted to a proportional treatment effect. This interconnection not only enables comparison of treatment effects across trials and endpoints but also offers new possibilities and flexibility in analyzing clinical trial data. For example, a shared proportional treatment effect can be used to model multiple endpoints with different scale ranges simultaneously, instead of modeling them sequentially. We hope that the enhanced capabilities of a proportional treatment effect to empower and connect, compared to the traditional test statistic of the difference between means, will attract the attention of statisticians so that this approach can be used more broadly.

There are some limitations to our study. First, the derivation is based on the delta method that is used to approximate the test statistic and might not be true for other approximation approaches. If the power gain is attributed to the delta method, our derivation reveals a potential limitation in commonly used statistical software packages when estimating nonlinear proportional treatment effects. It is crucial for statisticians to be aware of this issue. To the best of our knowledge, this matter has not been addressed in the existing literature. Second, although, in theory, a proportional treatment effect can yield more power than the difference between groups, it can become unstable when the mean change in the denominator (i.e., *μ*_*P*_) is close to zero, an issue inherent to any test statistic based on a ratio. Similar to Fieller’s theorem, which does not work well when the denominator is close to zero, we do not recommend the use of a proportional treatment effect under this circumstance either.^10^ To apply a proportional treatment effect, we recommend considering the following factors: (i) Whether a reasonably large placebo change over time is observed in natural history data or proof-of-concept phase I/II clinical trials, if available; and (ii) Conduct extensive simulations to evaluate the effectiveness and stability of a proportional treatment effect if the placebo change is small.

The availability of well-established statistical packages for estimating a proportional treatment effect and the theoretical demonstration of its potential to empower and connect hopefully provide an alternative test statistic for clinical trials with continuous, longitudinal endpoints.

## Data Availability

All data produced in the present work are contained in the manuscript and can be generated using the SAS codes provided in supplemental material.

## Disclosure of Conflict of Interests

Guoqiao Wang, PhD, is the biostatistics core co-leader for the DIAN-TU. He reports serving on a Data Safety Committee for Eli Lilly and Company, Amydis Corporate, Abata Therapeutics, and statistical consultant for Eisai inc. and Alector Inc.

The other authors report no COIs.

## Supplemental Materials

### Simulation Sample Code

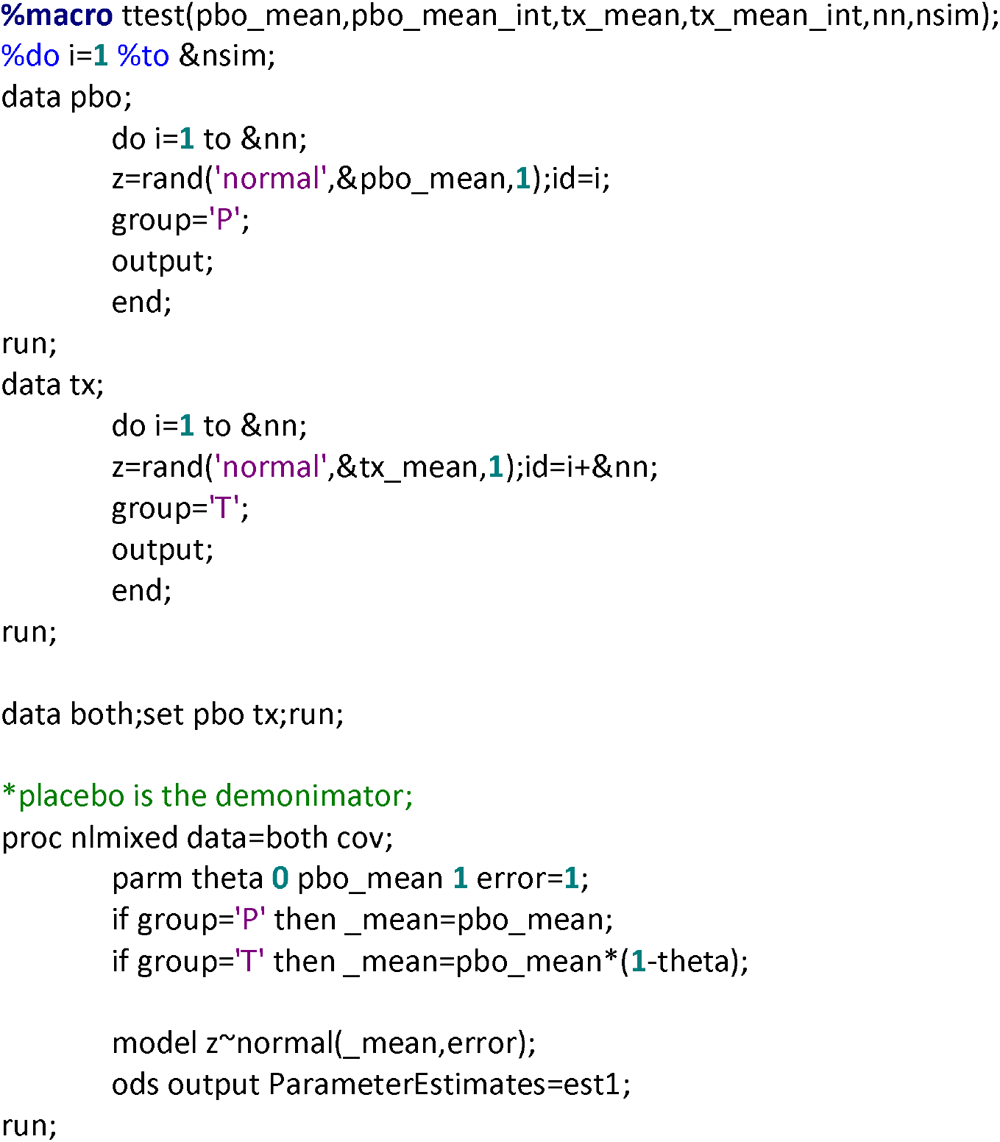
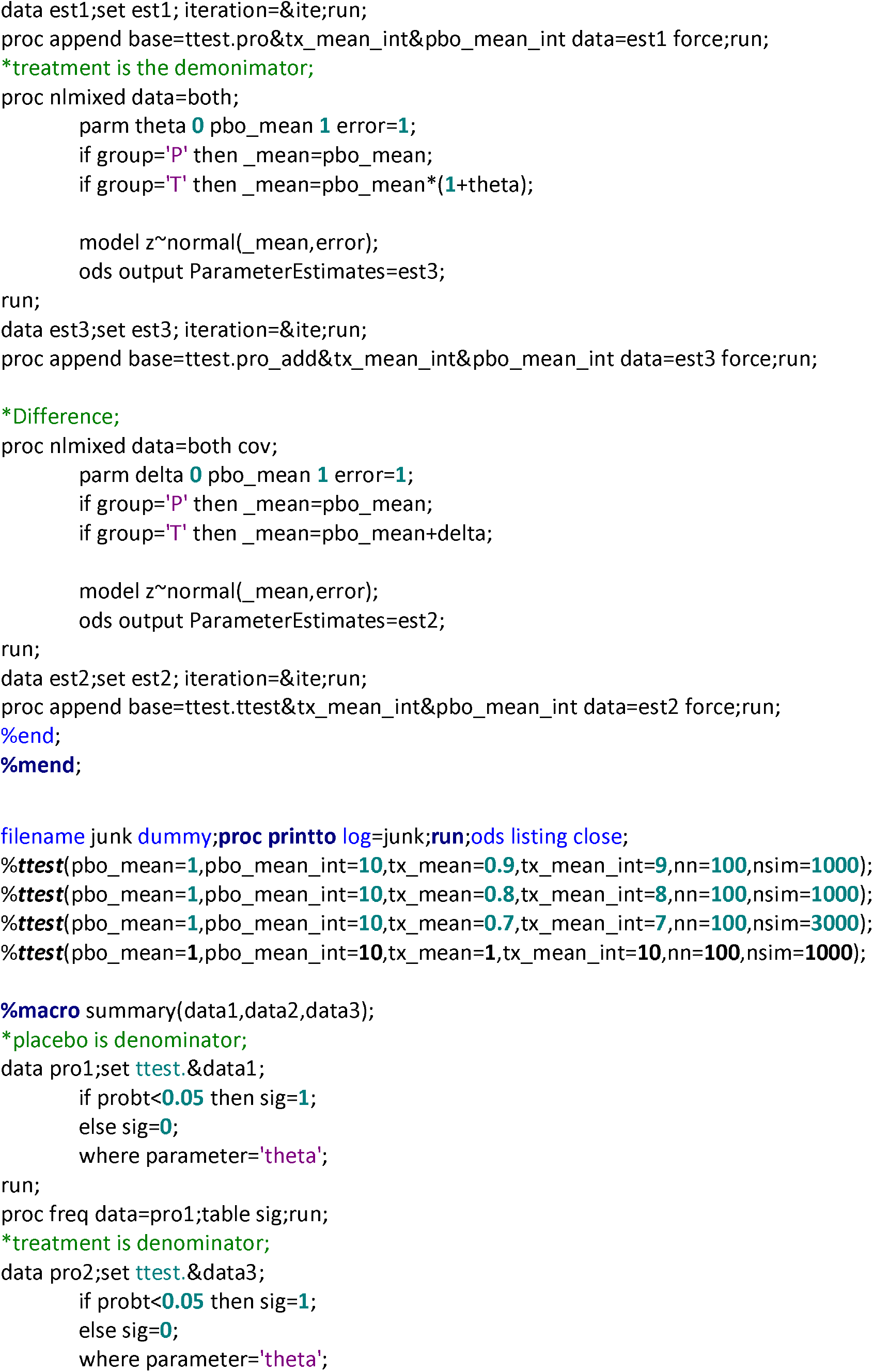
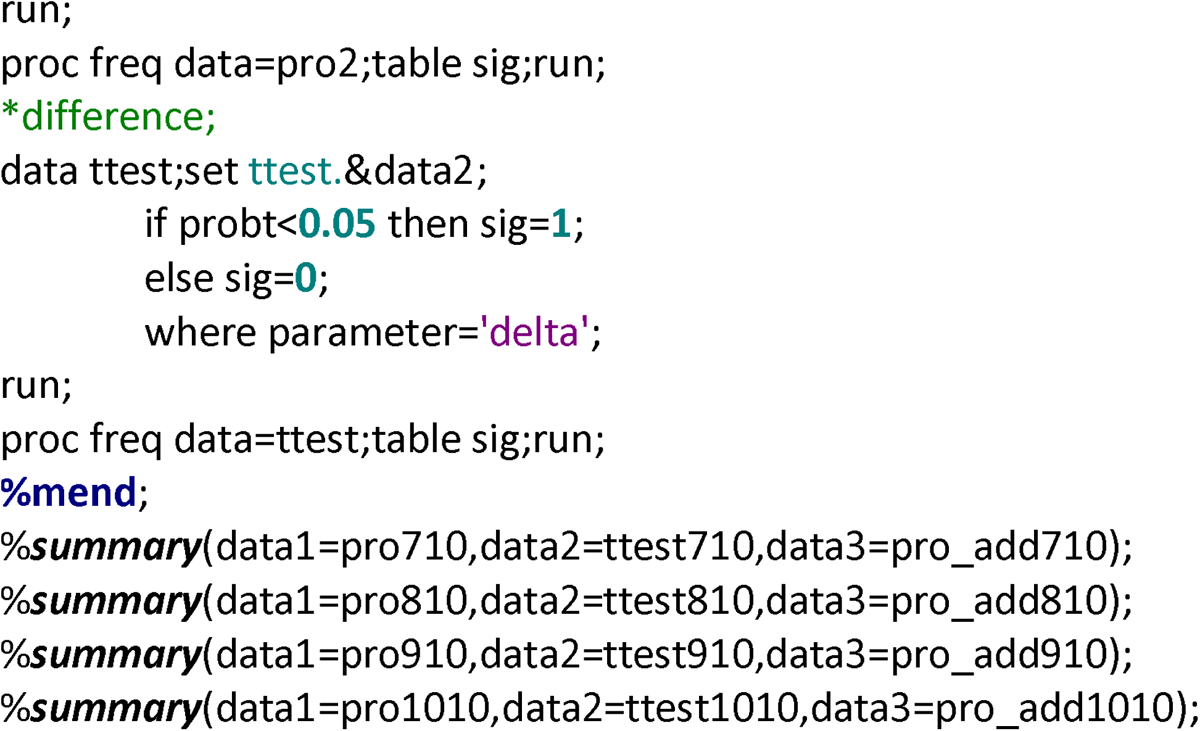

## Notes

### Funding Statement

This study did not receive any funding

